# Spatiotemporal Dynamics of Human Metapneumovirus and Potential Impact of Respiratory Syncytial Virus Interventions in the United States

**DOI:** 10.64898/2026.06.01.26354616

**Authors:** Ke Li, Stephanie Perniciaro, Jiye Kwon, Nathan D. Grubaugh, Daniel M. Weinberger, Virginia E. Pitzer

**Affiliations:** Department of Epidemiology of Microbial Diseases, Yale School of Public Health, New Haven, CT, USA; Public Health Modeling Unit, Yale School of Public Health, New Haven, Connecticut, USA; Department of Ecology and Evolutionary Biology, Yale University, New Haven, Connecticut, USA; Yale Institute for Global Health, Yale University, New Haven, Connecticut, USA

## Abstract

Human metapneumovirus (HMPV) causes acute lower respiratory infections, primarily affecting young children and older adults, with seasonal outbreaks peaking annually in March or April in the United States and other temperate regions in the Northern hemisphere. However, the factors driving HMPV seasonality in the United States remain poorly understood. We analyzed laboratory-confirmed HMPV cases and age-specific emergency department visits across 10 US regions, fitting an age-stratified dynamic transmission model to assess spatiotemporal patterns and investigate the influence of environmental variables and viral interference from RSV on HMPV transmission rates. We found that models incorporating climate variables into the transmission rate, including vapor pressure, precipitation, potential evapotranspiration, and minimum temperature, could not capture the timing of HMPV activity across all regions. Instead, HMPV timing was associated with RSV activity, with the HMPV transmission rate reduced in the presence of RSV. We showed that, unlike RSV, only models incorporating viral interference could reproduce the biennial pattern of HMPV observed in some regions, characterized by alternating “late-small” and “early-large” epidemics. Furthermore, our model successfully reproduced post-COVID-19 HMPV and RSV epidemics and predicted that RSV interventions are not likely to lead to a substantial increase in HMPV activity despite decreasing competition from RSV. Our work unravels the spatiotemporal dynamics of HMPV and its interaction with RSV, informing future seasonal forecasting and intervention strategies for HMPV.

**Author Summary:** Human metapneumovirus (HMPV) circulates each year in the United States and contributes to respiratory illness, particularly among young children and older adults. Although HMPV epidemics show clear seasonal patterns, the mechanisms underlying these patterns are not well understood. In this study, we combined surveillance data from multiple regions in the United States with mathematical modeling to investigate the drivers of HMPV transmission. We evaluated whether environmental factors or interactions with respiratory syncytial virus (RSV) better explained differences in epidemic timing and intensity. Our findings indicate that interactions between HMPV and RSV play an important role in shaping HMPV epidemics. In particular, RSV circulation appeared to suppress HMPV transmission, helping explain the alternating epidemic patterns observed in some regions. Our model also suggested that expanded RSV prevention programs are unlikely to substantially increase HMPV burden. This work provides new insights into respiratory virus interactions and highlights the importance of considering pathogen interactions when predicting seasonal outbreaks and evaluating intervention strategies.

## Introduction

Human metapneumovirus (HMPV) is a major cause of acute respiratory infections globally, leading to significant hospitalizations and deaths, particularly among young children, older adults, and other high-risk groups. In children, its disease burden is similar to that of influenza and respiratory syncytial virus (RSV), while in older adults, HMPV contributes substantially to morbidity and healthcare utilization [1,2]. The reported incidence of HMPV varies across age groups and regions, with most data based on medically attended cases. In 2018, nearly 650,000 children under 5 years were hospitalized with HMPV-related acute lower respiratory tract infection globally, resulting in over 7,500 in-hospital deaths [1]. Data on the burden of HMPV in adults are limited; there were 12.1 hospitalizations and 0.3 deaths per 100,000 adults in the United States (US) between 2016 and 2019 [3]. These reported numbers likely underestimate the true burden, due to limited testing and underreporting of HMPV infections. Despite its global impact, no licensed vaccines or antivirals against HMPV are available, and surveillance remains limited, particularly in resource-constrained settings [4].

HMPV demonstrates distinct seasonal patterns in most geographic locations, influenced by latitude and climate [5]. Before the COVID-19 pandemic, HMPV typically circulated annually, peaking in March or April in the United States and other temperate settings [6–8]. Tropical and subtropical areas have less pronounced HMPV seasonal patterns, with increased detections frequently observed during the more humid wet seasons, but with high variations in timing between years [9,10].

Gaps remain in our understanding of the relative impact and interplay of determinants of HMPV transmission. Addressing these gaps is crucial for several reasons. First, it will inform the development of public health strategies aimed at preventing and controlling seasonal viral transmission. Understanding and predicting the seasonality of HMPV is essential in determining the optimal timing of immunoprophylaxis. Second, it will guide future studies in assessing and quantifying the impact of these strategies and help predict how interventions against other viruses could influence HMPV. Third, it will support the creation and implementation of practical predictive tools to forecast seasonal HMPV activity.

Previous studies have shown that a higher amplitude of seasonal forcing, associated with climatic factors, could explain the seasonality of RSV in the US, in particular the biennial epidemics observed in the West and upper Midwest [11]. Studies examining the association between climatic factors and HMPV seasonality, however, have demonstrated inconsistent findings, likely due to variations in analytical approaches and differences across regions or study sites [12–17]. Besides climatic factors, viral interference—defined as the phenomenon whereby infection with one virus alters the likelihood or severity of infection with another—between HMPV and RSV has been observed at both the host and population levels, influencing the dynamics of HMPV transmission [18–21]. Mathematical modeling of disease transmission dynamics provides a powerful approach to explore the mechanistic relationships between potentially important environmental variables and seasonal variation in transmission rates [11,22–26]. Through such models, we can better understand how environmental and other factors may influence the transmission dynamics of different respiratory viruses.

In this work, we began by analyzing weekly laboratory-confirmed positive tests for HMPV from 2008 to 2025 across 10 regions of the US, describing the spatiotemporal patterns of HMPV activity. We then constructed a mechanistic, age-stratified mathematical model to investigate potential drivers of HMPV transmission. Using a maximum likelihood approach, we estimated climatic and viral interference parameters and compared different models based on our hypotheses. Finally, we applied the best-fit model to predict RSV dynamics under various RSV intervention scenarios and assessed the impact on HMPV burden, exploring how changes in RSV epidemics could influence the timing and intensity of HMPV activity.

## Results

### Epidemiology and clinical impact of HMPV

HMPV exhibits distinct seasonal patterns across the 10 different administrative regions of the US Department of Health and Human Services (HHS). We observed that the number of positive laboratory tests for HMPV from July 2008 to June 2025 was strongly seasonal, showing an annual pattern of outbreaks during the late winter/early spring months in the US (**Fig. 1A, Fig. S1**). Some regions (e.g., regions 8 and 9) exhibited biennial patterns, with alternating “late-small” epidemics in even-numbered years (e.g., 2010/11 season) and “early-large” epidemics in odd-numbered years (e.g., 2011/12 season). HMPV activity was disrupted by the COVID-19 pandemic during the 2020/21 season, with activity gradually returning to normal in the 2023/24 and 2024/25 seasons. Year-to-year variations in positive test counts likely reflect increasing testing frequency. To allow consistent seasonal comparisons, we adjusted HMPV-positive counts to account for changes in testing practices, ensuring that observed trends better represent underlying HMPV activity rather than variations in testing intensity. See **Materials and Methods** for details.

**Figure 1.**
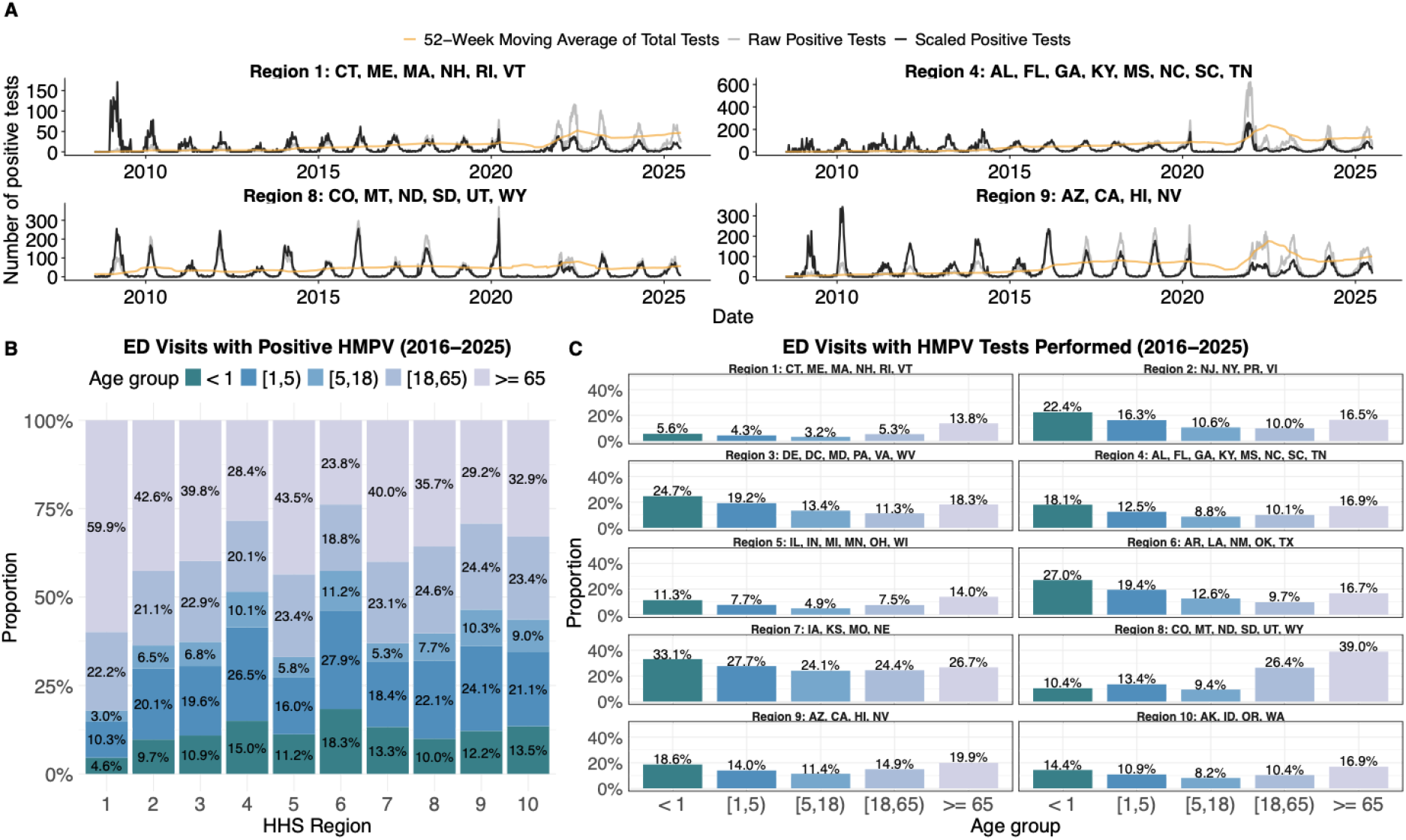
Seasonal patterns and age distributions of HMPV cases in the 10 different administrative regions of the US Department of Health and Human Services. **(A)** Time series of weekly positive laboratory tests for HMPV in Region 1, Region 4, Region 8 and Region 9. Raw data is shown in grey, while the scaled data accounting for the change of testing frequency (orange curve) over time is shown in black. Time series of other regions are shown in **Fig. S1**. Age distribution of **(B)** ED visits positive for HMPV, and **(C)** ED visits with HMPV tests performed across 10 regions. The states and their abbreviations for each HHS region are listed in **Table S1**.

We further stratified the total number of emergency department (ED) visits with a positive HMPV test into five age groups (**Fig. 1B, Fig. S2**). Across different HMPV seasons in the US, children younger than 1 year and those aged 1–5 years accounted for an average of 11.9% and 20.6% of the cases, respectively. Adults above 65 years represented the largest proportion of cases, averaging 37.6% across the 10 regions. Children and adults aged 5–18 years and 18–65 years accounted for 7.6% and 22.4% of the cases, respectively. In addition, we observed regional variations in the age distribution of HMPV-positive tests. This is likely due to differences in testing frequency and practices in different regions (**Fig. 1C**). For example, on average, 5.6% of ED visits for infants under 1 year old and 13.8% of visits for older adults aged 65 years and above were tested for HMPV in Region 1 from 2016 to 2025. By contrast, the corresponding percentages were 22.4% and 16.5% for the same age groups in Region 2.

### Description of spatiotemporal patterns of HMPV

We next sought to quantify the timing of HMPV epidemics in the US. To do this, we computed the center of gravity of each HMPV season in each region (see **Materials and Methods**). Laboratory reports from the 10 HHS regions of HMPV-positive specimens exhibited a distinct spatial pattern, with mean timing of HMPV activity from 2008 to 2019 (before the COVID-19 pandemic) occurring earliest in the southeastern US (region 4) in early February and latest in the upper Midwest (region 8) in early March (**Fig. 2A**). Following the COVID-19 pandemic, HMPV had an earlier epidemic in 2021/22, beginning in early November in regions 4 and 6 and ending in late March in region 10. The spatiotemporal pattern of HMPV activity gradually returned to normal from 2023 onwards.

**Figure 2.**
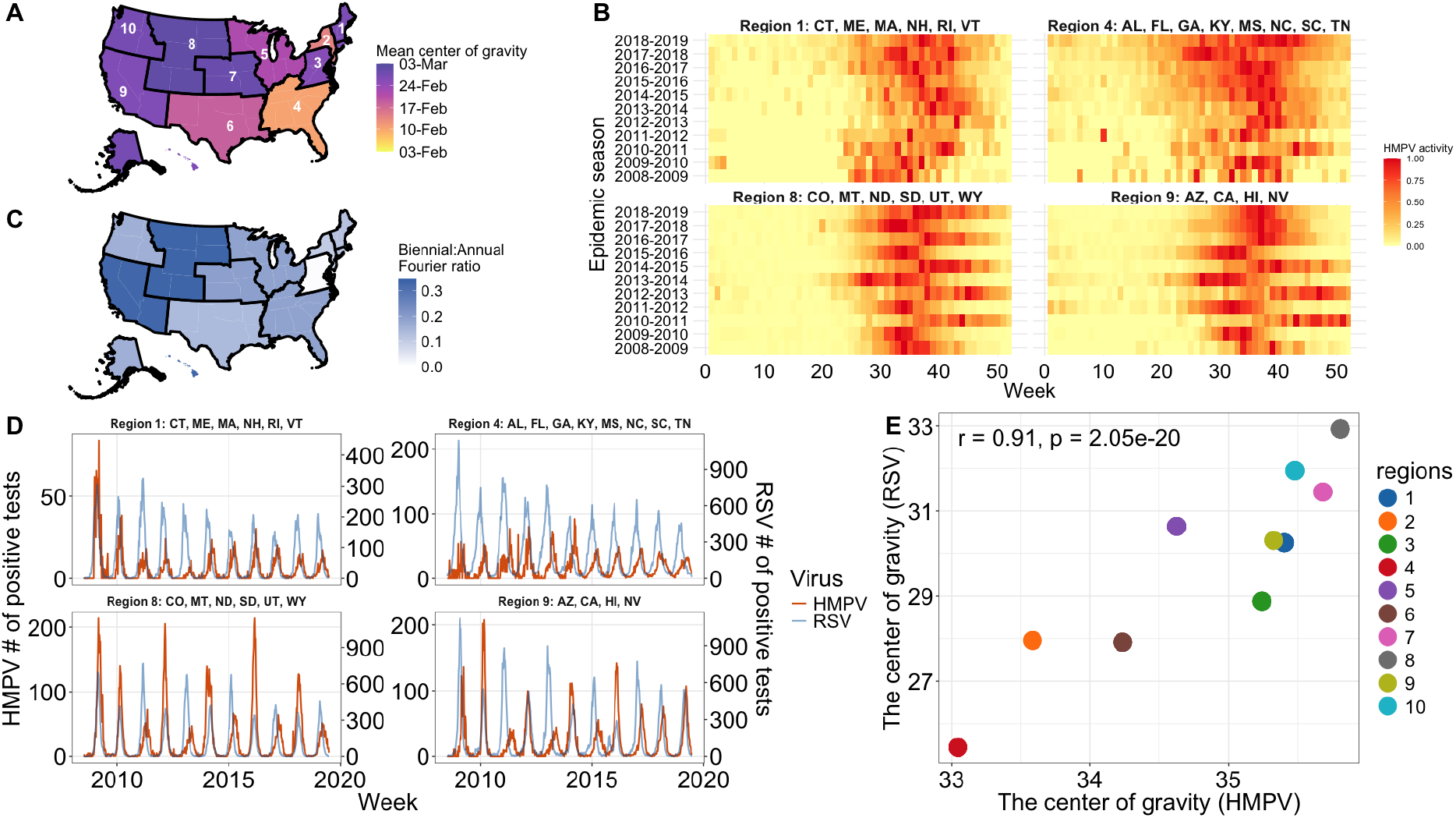
Spatiotemporal patterns of HMPV activity in the United States. **(A)** Center of gravity of HMPV activity in the US before the COVID-19 pandemic. **(B)** Heatmaps of HMPV epidemics before the COVID-19 pandemic in select regions. HMPV activity was normalized by region and season to a 0–1 scale, using positive laboratory test results adjusted for testing frequency. An epidemic season is defined as starting from July of one year (i.e., week = 1) to June of the following year (i.e., week = 52). **(C)** Strength of biennial cycle in HMPV activity, as indicated by the ratio of the biennial to annual Fourier amplitude. (**D**) Time series of weekly positive laboratory tests for RSV (blue curve) and HMPV (red curve) for select regions. The number of positive cases are adjusted for the total number of tests. RSV cases were scaled by dividing by 2 to make their magnitude comparable to that of HMPV for visual comparison. (**E**) The correlation between the mean timing of HMPV and RSV epidemics in 10 regions from 2008 to 2019.

Within regions, the seasonal timing of HMPV circulation remained relatively stable from year to year (**Fig. 2B, Fig. S3**). While most regions exhibited annual peaks (e.g., regions 1 and 4), some showed a biennial pattern of HMPV epidemics (e.g., regions 8 and 9); these regions were concentrated in the upper Midwest and western United States (**Fig. 2C**), with epidemics occurring in early February during odd-numbered seasons and in mid-March during even-numbered seasons. Notably, we observed alternating biennial epidemics of HMPV and RSV in some US regions, with small HMPV epidemics coinciding with large RSV seasons (regions 8 and 9, **Fig. 2D, Fig. S4**), and an overall pattern for RSV and HMPV epidemic magnitudes to be negatively correlated across regions, although the strength of this relationship varied by region. The inverse association was most apparent in the regions exhibiting biennial activity, whereas others (e.g., regions 1, 2, and 10) showed weak or minimal correlations (**Fig. S5**). This variability across regions could be explained by regional differences in population structure or the timing of seasonal circulation. Nevertheless, there was a strong positive correlation between the mean timing of RSV and HMPV epidemics across all 10 regions (Pearson correlation coefficient *r* = 0.91, *p* < 0.05; **Fig. 2E**). The result suggests that the seasonality of HMPV may be closely linked to RSV activity, potentially reflecting interactions between the two viruses.

### Dynamic model analyses

Having demonstrated the spatiotemporal patterns of HMPV activity in the US, we next sought to explore the mechanistic relationships between seasonal variation in the transmission rate and potential drivers. We developed an age-stratified SIS (Susceptible-Infectious-Susceptible) model of HMPV transmission dynamics (**Fig. 3A, Eq. (1)**), accounting for repeat infections and incorporating natural history parameters derived from observational studies (**Table S2**). See **Materials and Methods** for a detailed model description. We fitted the model to both weekly HMPV-positive counts from 2008 to 2019 (i.e., pre-COVID-19 seasons) and the age distribution of ED visits (aggregated from 2016 to 2025), estimating key transmission parameters. A similar model was previously developed and fitted to data on RSV [11].

**Figure 3.**
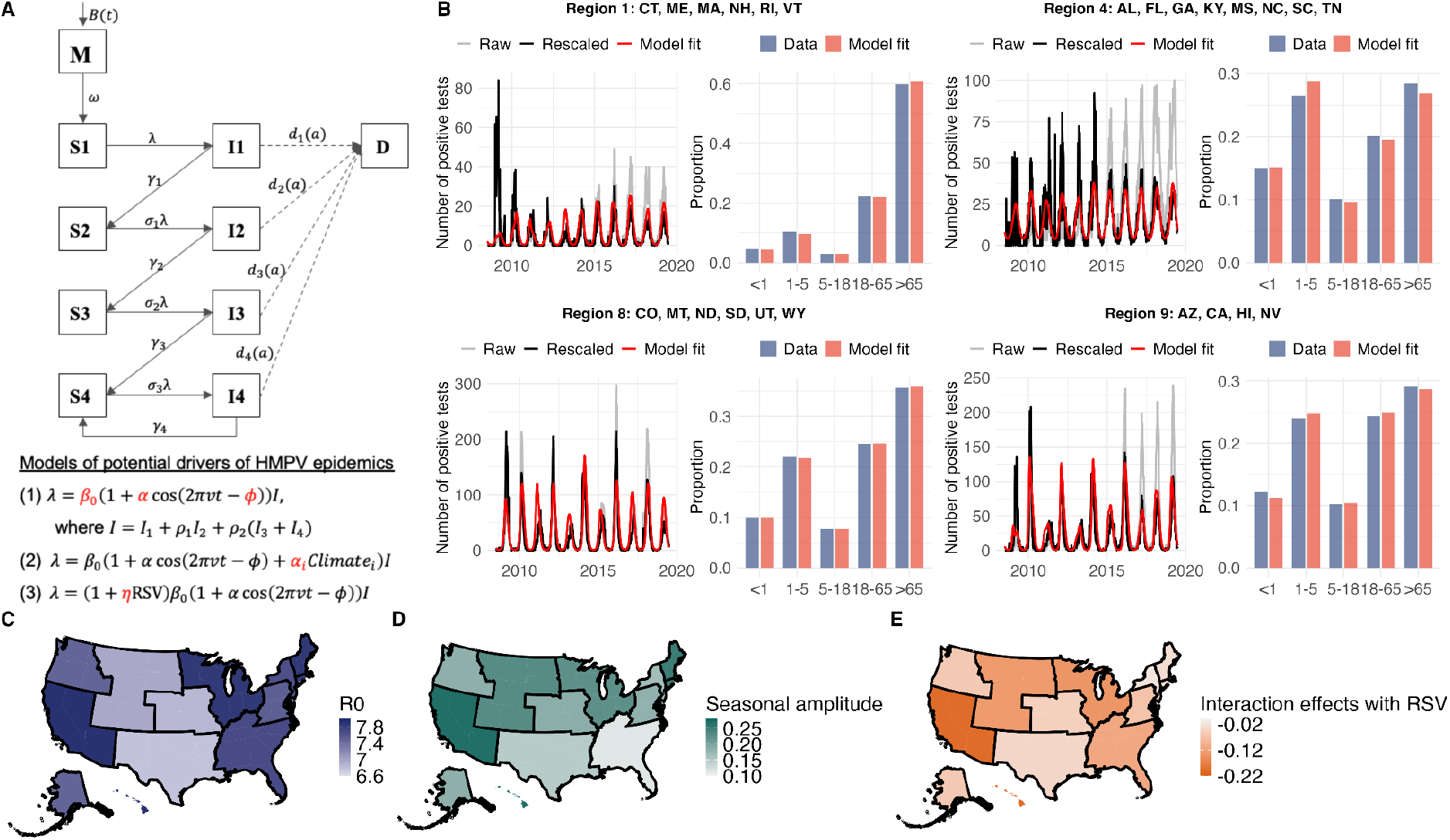
Fit of dynamic transmission model for HMPV. **(A)** Model diagram illustrating the structure of the HMPV transmission model (age is omitted for simplicity). Compartments *S1-S4* represent susceptible populations, and compartments *I1-I4* represent infected populations. Infants are born at rate *B(t)* into the population with maternal antibodies that protect them from infection and disease, represented by *M*. Compartment *D* represents the population that develops severe lower respiratory infections; note that the compartment represents an observed state rather than infection state in the model. See **S1 Text** for detailed model equations. **(B)** Model fits to weekly HMPV positive tests, and age distributions of HMPV ED visits for select regions. Raw data are shown in grey, while the scaled data accounting for the change of testing frequency are shown in black. The best model predictions are shown in red. Model fits for other regions are shown in **Fig. S9**. Parameter estimates of the region-specific **(C)** basic reproduction number, **(D)** seasonal amplitude, and **(E)** interaction effects with RSV.

We investigated multiple climate variables—including temperature, vapor pressure, precipitation, and potential evapotranspiration (PET)—as candidate drivers of transmission rates, since environmental conditions have been shown to influence the spread of other respiratory virus epidemics, including RSV [11]. In addition, we hypothesized that RSV viral interference may influence HMPV transmission, as HMPV epidemics in temperate settings typically follow RSV peaks [5,27], and we observed alternating biennial RSV–HMPV epidemics in some US regions. We considered two competing hypotheses for the observed biennial seasonality of HMPV in some regions. One hypothesis is that the alternating epidemic patterns arise from shared environmental or climatic drivers that similarly affect both RSV and HMPV transmission. An alternative hypothesis is that RSV directly alters HMPV transmission dynamics through viral interference. These mechanisms are not mutually exclusive, and both may contribute to the observed epidemic patterns.

To assess the impact of climate variables on HMPV transmission, we incorporated normalized time-series of climatic data into the model, assuming that transmission rates varied in proportion to these factors (**Fig. 3A, Eq. (2)**). To investigate whether viral interference from RSV influences HMPV transmission, we assumed that RSV incidence modifies HMPV transmission rates by a multiplicative factor, η, consistent with previous modeling studies [18]. Normalized weekly RSV-positive tests were used directly as an input, with the multiplicative effect proportional to RSV case counts (**Fig. 3A, Eq. (3)**). We again fitted the model to the same data to estimate both key transmission and climate-related or viral interference parameters.

We found that, among all variables tested in our analysis, only viral interference significantly impacted HMPV transmission rates and improved model fit as measured by AIC (**Fig. S6-S9, Table S3**). The best-fitting model (with viral interference) was able to reproduce the seasonal pattern of RSV positive tests and the age distribution in all 10 regions (**Fig. 3B, Fig. S10**), with the correlation between observed and predicted center of gravity *r =* 0.85 (*p* = 0.002, **Fig. S11**). Notably, we found that only the model with viral interference effects from RSV was able to reproduce the biennial pattern of epidemics observed in regions 8 and 9. We also simultaneously assessed the impact of both climatic variables and viral interference effects from RSV on HMPV transmission. We found that this combined model did not perform better than the model including only the viral interference effect (**Fig. S12, Table S3**). Although incorporating climate variables directly into the model did not improve model fit, we observed negative correlations between mean precipitation, mean vapor pressure and estimated seasonal amplitude (**Fig. S13**).

Based on parameter estimates, the mean basic reproduction number (R_0_) of HMPV was estimated at 7.5, with region-specific values ranging from 6.8 to 7.9 (**Fig. 3C**). The mean amplitude of seasonal forcing was 0.17 (range: 0.11–0.27; **Fig. 3D**), with higher amplitudes observed in regions with biennial HMPV epidemics. Unlike with RSV, the higher seasonal forcing alone could not reproduce the biennial epidemics of HMPV in these regions (probably due to the lower R_0_). A value of 0.17 implies that the transmission rate at its seasonal peak is approximately 17% higher than the average annual baseline. Parameter estimates indicated a suppressive effect of RSV incidence on HMPV transmission, with a mean of −0.08 across all 10 regions (range from −0.2 to −0.03; **Fig. 3E**). The mean value implies that, at peak RSV seasonal activity—when normalized RSV incidence reaches its maximum—HMPV transmissibility is reduced by 8% relative to weeks with minimal RSV co-circulation. We also used the profile likelihood method (**see Materials and Methods**) to compute a 95% confidence interval (CI) for the interaction parameter in each region (**Table S4**). We found that the effect is statistically distinguishable from zero, indicating a robust signal of viral interference across all regions.

### Predictions of HMPV transmission dynamics under RSV interventions

Having demonstrated a suppressive viral interference effect of RSV on HMPV transmission rates, we next assessed how RSV interventions might alter HMPV epidemics. To do this, we calibrated the same transmission model to RSV-positive tests from 2008–2019 and reproduced pre-COVID-19 epidemics for all 10 regions (**Fig. 4A**, shaded red; **Fig. S14**). We then ran forward simulations incorporating Google mobility data (see **Materials and Methods**) to adjust RSV transmission during 2020–2025 (**Fig. 4A**, shaded yellow; **Fig. S14**). We then projected the influence of different RSV interventions on both RSV and HMPV cases from 2025 to 2027. Although the model did not capture the first disrupted season following the pandemic, it reproduced RSV epidemics from 2023 onward. The model-predicted RSV cases for 2008–2019 were also able to reproduce the pattern of HMPV cases in the model with interference (**Fig. S15**). Using the same method, we showed that forward simulations for HMPV successfully predicted post-pandemic epidemics (**Fig. 4B**, shaded yellow; **Fig. S16**).

**Figure 4.**
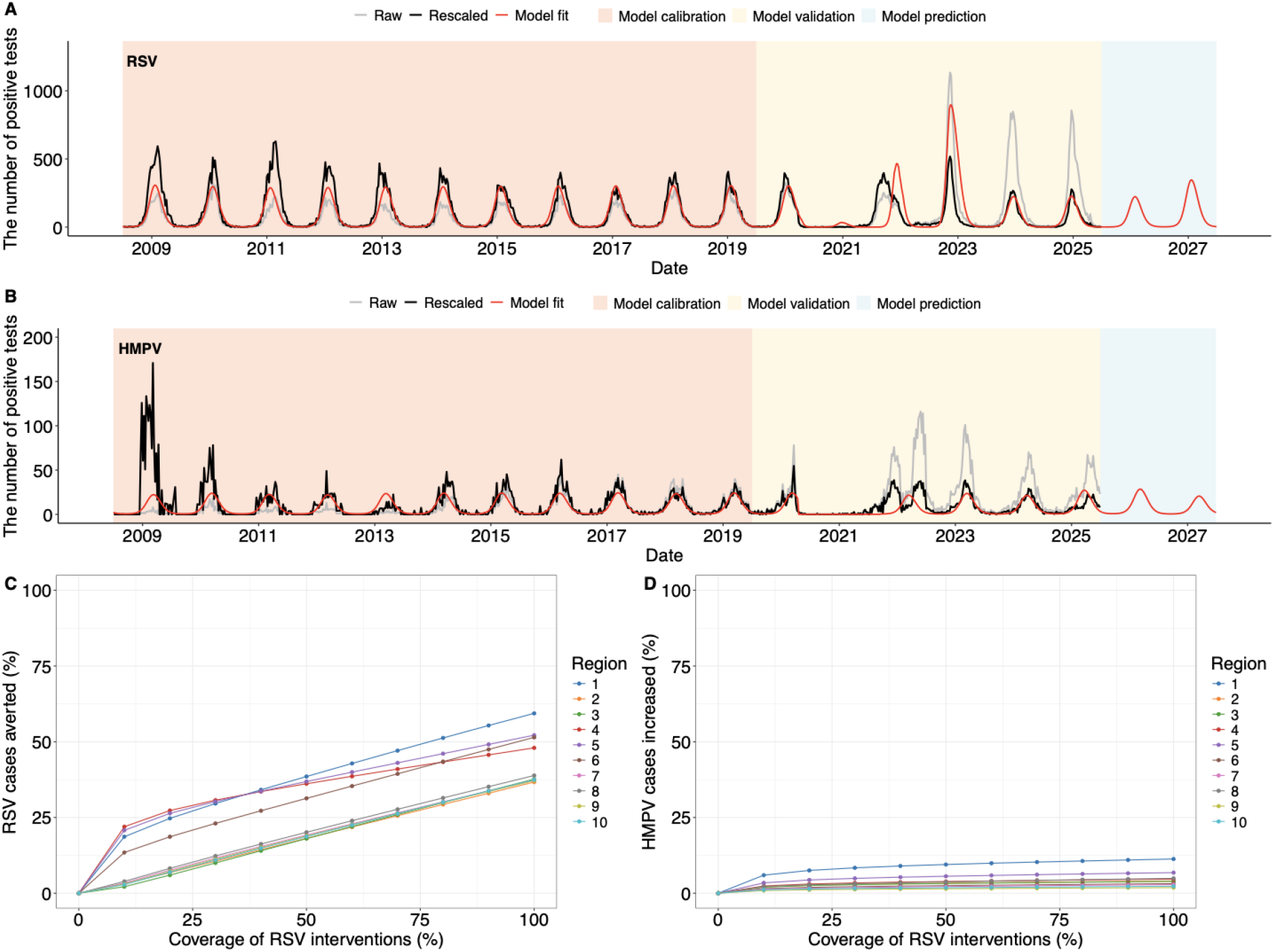
Model validation and prediction of epidemics and HMPV burden under RSV interventions. Model fits to **(A)** weekly RSV-positive tests, or **(B)** weekly HMPV-positive tests from 2008 to 2019 for Region 1 are shown in red (shaded red area). Model validation incorporating Google mobility data to modify transmission rates from 2019 to 2025 is shown in yellow, and model predictions under RSV interventions are shown in blue. Raw data are shown in grey, and scaled data (accounting for changes in testing frequency) are shown in black. Other regions are given in **Fig. S11-12**. The relationship between RSV interventions coverage (%) and **(C)** the decrease (%) of RSV-related LRT infections, and **(D)** the increase (%) of HMPV-related LRT infections during 2025-2027.

To simulate RSV intervention scenarios, we extended the model to include three intervention strategies: 1) monoclonal antibody protection against RSV for newborns, 2) maternal immunization, and 3) vaccination for older adults aged 65 and above (**Fig. S17**). We assume that coverage begins in the 2023/24 season. We then simulated RSV-related lower respiratory tract (LRT) infections under coverage rates from 0% to 100% and assessed the corresponding reductions in RSV infections starting in 2025 (**Fig. 4A**, shaded blue; **Fig. S14**). Throughout the simulations, the proportion of older individuals vaccinated is assumed equal to the sum of the proportions of vaccinated newborns and mothers, with newborn and maternal vaccination proportions being equal. The simulations estimated that RSV-related LRT cases would decline by 25.7% in the overall population at 50% coverage of all interventions (range across the 10 regions: 21.3%-33.9%; **Fig. 4C**). Reductions were larger for infants aged <1 year: 40.3% (37.1%–44.5%), and for older adults aged above 65: 55.7% (52.4%–61.1%; **Fig. S18**). Regional differences likely reflect variation in baseline RSV burden and demographic structure.

We also conducted sensitivity analyses assuming that only one intervention was used. We found that implementing monoclonal antibodies alone could avert the infections by 36.9% (31.5%–40.2%) and 1.43% (0.84%–1.68%) in infants under 1 and older adults, respectively, at the same coverage level (**Fig. S19**). Implementing maternal vaccination alone could reduce RSV-related LRT infections by 31.3% (26.5%–34.4%) in infants under 1 year and by 0.3% (0.11%–0.37%) in older adults at 50% coverage (**Fig. S20**). If vaccination for older adults alone were implemented at 50% coverage, we predicted that it could reduce RSV-related LRT infections by 8.9% (5.7%–15.3%) in infants under 1 year and by 51.5% (48.5%–55.9%) in older adults, respectively (**Fig. S21**).

By contrast, we predicted that the reduction of RSV cases (at 50% coverage) during 2025-2027 would have a negligible effect on HMPV epidemics with a mean 2.01% increase in overall HMPV-related LRT infections (0.80%–3.22%; **Fig. 4D**), similarly minimal among infants aged between 0-24 months and older adults (**Fig. S22**). We also predicted that the implementation of RSV interventions would not change the timing of RSV or HMPV epidemics (**Fig. S23**). We further conducted additional sensitivity analyses. We varied the parameters associated with the RSV–HMPV interaction and predicted HMPV incidence under RSV intervention scenarios in each region. We found that the result remains qualitatively consistent (**Fig. S24**).

## Discussion

In this work, we investigated the spatiotemporal pattern of HMPV activity in the US by analyzing longitudinal laboratory-confirmed HMPV tests from 2008-2025. Using an age-stratified dynamic transmission model, we evaluated multiple drivers for the observed seasonal and biennial variation of HMPV epidemics, which would help build predictive models of HMPV incidence. We showed that incorporating viral interference effects from RSV significantly improved the model fit and could replicate the biennial HMPV pattern in certain regions, whereas climatic factors could not explain the seasonal patterns of HMPV. Identifying viral interference from RSV on HMPV epidemics could help evaluate whether the rollout of RSV interventions would shift or alter HMPV epidemics, potentially increasing disease burden. We demonstrated that although RSV shows a suppressive viral interference effect on HMPV, the overall burden of HMPV is not expected to increase in the coming epidemic seasons with the continued rollout of RSV interventions.

Although our study addresses questions related to those examined by others [18], it provides several distinct contributions. First, we are the first study to analyze and characterize HMPV epidemics in the US, demonstrating the spatiotemporal patterns of HMPV activity across different regions and identifying annual and biennial patterns of activity. Second, our age-stratified modeling framework captures key differences in susceptibility, contact patterns, and disease burden across age groups, which are central to understanding HMPV transmission. Third, by adopting a model structure parallel to prior RSV analyses [11], we enable direct, side-by-side comparison within a unified framework using the data in the US for both RSV and HMPV. Our results also emphasize that the mechanism is not sensitive to the underlying structural assumptions, as our model structure differs from that in [18]. Lastly, the age-stratified structure allows us to simulate age-targeted RSV intervention scenarios and quantify their downstream impact on HMPV dynamics, generating public health insights that extend beyond previous aggregate models.

HMPV exhibits a consistent seasonal pattern in the US, with epidemics typically peaking in late winter to early spring and displaying regional variations. This is in line with prior surveillance studies in the US [28–30]. Compared to RSV, HMPV displays similar spatial gradients across regions, beginning in the Southeast, moving through the Northeast and Midwest, and eventually ending in the Southwest and West [11,31]. However, HMPV peaks were closer across regions, with most areas peaking within about a month of each other, whereas RSV showed clearer spatial differences, typically starting in the Southeast in late December and ending in the West by February. Similar to previous findings for RSV [11], we also observed that greater variability in climatic factors was associated with higher seasonal amplitudes, although incorporating climate variables directly into the model did not substantially improve model fit—likely because the timing of climatic fluctuations did not fully align with epidemic peaks. Thus, while RSV interference appears to influence HMPV dynamics, climatic factors may also contribute either directly or indirectly through their influence on RSV.

Our findings suggest that, among all variables tested, HMPV transmission is less sensitive to climate factors. This may help explain the smaller regional variation in its seasonal timing compared to RSV. However, we cannot exclude the possibility that other environmental drivers also contribute to HMPV transmission dynamics. Furthermore, both HMPV and RSV exhibit similar biennial patterns across regions. However, the timing and intensity of these patterns alternated between the two viruses. This pattern may provide important supportive evidence that the circulation of respiratory viruses is not isolated but instead shaped by interactions among the co-circulating pathogens. Recognizing this interplay is critical for understanding seasonal dynamics and for anticipating future epidemic behavior. These findings highlight both the similarities and differences in the spatiotemporal dynamics of HMPV and RSV, underscoring the need to account for virus-specific drivers when interpreting patterns of seasonal epidemics.

Studies examining the statistical association between different climatic variables and HMPV seasonality have reported mixed results [12–17]. Some analyses found associations between HMPV activity and meteorological factors such as temperature, humidity, or rainfall, while others did not detect significant relationships. Overall, these findings suggest that climatic conditions may influence HMPV transmission in certain geographic locations, but the strength and consistency of these associations vary. In temperate regions, HMPV seasonality appears only weakly linked to climate, or the influence of climatic conditions on HMPV transmission may instead be indirect and mediated through their effect on RSV. Mechanistic models provide a valuable framework for disentangling causal relationships, integrating heterogeneous mechanisms, and testing alternative hypotheses. Our analysis showed that variations in vapor pressure, minimum temperature, precipitation, and PET did not substantially influence HMPV transmission rates. One possible explanation is that the regional scale of analysis may lack the spatial resolution needed to identify localized climatic effects influencing transmission dynamics. Another explanation is that climate variables may lack independent explanatory power because their effects on transmission could operate indirectly through behavioral changes. For example, temperature may influence the amount of time individuals spend indoors, thereby increasing close-contact interactions that facilitate HMPV transmission. Because these behavioral responses are also correlated with seasonal changes in the HMPV transmission rate disentangling the direct effects of climate from those of behavior in the analysis is challenging.

The differences in climate sensitivity between RSV and HMPV may be explained by several epidemiological and virological factors. First, HMPV exhibits a moderate level of environmental stability. Studies have shown that HMPV can remain infectious at ambient room temperature and 4°C, but its infectivity decreases at higher temperatures, which may limit its transmission during warmer periods [32]. In contrast, RSV has been observed to survive longer on surfaces and in aerosols under specific humidity and temperature conditions, making it more responsive to climatic fluctuations [33]. Second, although both HMPV and RSV spread through direct contact, droplets, and aerosols, the relative importance of these transmission routes may differ between the two viruses [34,35]. These differences suggest that HMPV dynamics are more strongly shaped by factors other than climate.

The presence of viral interactions between different respiratory viruses has been evident at the host level [22,36–40]. In particular, both within-host [21] and population-level [18–20] effects of viral interference between RSV and HMPV have been demonstrated in previous studies. One key finding from our model estimates is that a suppressive viral interference effect from RSV was identified as a factor influencing HMPV transmission rates in the US in the best-fitting model. Note that we did not explicitly model RSV epidemics within the HMPV transmission framework; instead, we incorporated observed RSV incidence directly into the force of infection to inform HMPV transmission rates. We acknowledge that viral interference can influence transmission through multiple pathways that our single-pathogen model could not capture. For example, host susceptibility to secondary infection may be impacted if an initial infection temporarily boosts immune defenses that protect against subsequent pathogens [41,42]. It can also reduce or enhance transmissibility during co-infection, as interactions between co-circulating viruses may suppress or amplify viral shedding, affecting how easily a host transmits one or both pathogens to others [43,44]. Interference may also shape post-infection outcomes by modulating immune responses or shifting viral replication and clearance dynamics [19,45–47]. These effects are not necessarily symmetric or unidirectional and can vary by context, such as household transmission [48]. While our model can detect deviations in transmission consistent with interference, it cannot resolve causal mechanisms. As more granular co-infection data become available, multi-pathogen models will be needed to capture the full spectrum of viral interference.

Our findings on RSV–HMPV interactions highlight the challenges of simplifying complex biological processes into a single parameter. While this parameter captures overall interference effects, it does not fully reflect the nuances of immunological protection or its temporal dynamics. As a result, the duration and intensity of viral interference remain difficult to disentangle. Unlike influenza–RSV interactions, which have been shown to be mediated primarily by innate immune responses and are therefore thought to be transient in nature [41,46], RSV–HMPV interactions may involve adaptive immune mechanisms that extend interference over longer time scales. For instance, recent structural work demonstrated that a monoclonal antibody (RSV-199) can cross-neutralize both RSV and HMPV through recognition of a conserved epitope on the viral fusion protein, suggesting a role for adaptive immunity in mediating cross-protection [49]. This raises the possibility that RSV–HMPV interference could persist beyond the short-lived effects typically attributed to innate immunity, although the precise duration and clinical significance of this protection remain uncertain. Altogether, these results underscore the need for future studies to elucidate the underlying biological mechanisms, which would allow for the development of more detailed models that explicitly incorporate these processes.

In the US, the extended half-life monoclonal antibody nirsevimab was approved in July 2023, followed by clesrovimab in June 2025—both intended for passive immunoprophylaxis of infants against RSV. During the 2023–24 RSV season, approximately 29% of infants received protection through either nirsevimab administration or maternal RSV vaccination, with coverage rates ranging from about 11% to 53% across states [50]. The continued use of monoclonal antibodies has proven effective in providing protection for infants [51–54], and our predicted reduction of RSV-related LRT infections in the pediatric group is consistent with previous observational studies [55]. Maternal RSV vaccination was introduced in late 2023 and reached an estimated 15–20% uptake among pregnant women in the first season, contributing substantially to infant protection [56]. For older adults, two RSV vaccines were approved in 2023, with national coverage among adults aged over 60 years estimated at roughly 20–25% during the first season of rollout [57], and they have been shown to be effective in reducing LRT infections [58,59]. Based on the inferred interactions between RSV and HMPV, we evaluated the potential for an increased HMPV burden following the introduction and continued use of RSV interventions in the coming seasons. We focused on the 2025–2027 horizon to capture the near-term impact of RSV immunization programs at their initial coverage levels, when projections are most reliable, while acknowledging that longer-term shifts in disease burden may require additional assumptions and carry greater uncertainty.

Our model predicted that RSV-related lower respiratory tract infections would be reduced following the interventions, while the overall HMPV burden—including in children aged 0–24 months and older adults >65 years—was unlikely to increase significantly. This finding is consistent with previous studies, which predicted that HMPV burden would not exceed levels observed during pre-intervention RSV epidemics [18]. Our model shows a modest impact of RSV immunization on HMPV burden, which can be explained by the following two reasons. The estimated interference parameter should be interpreted as the maximum instantaneous suppression of HMPV transmissibility during periods of peak RSV circulation rather than as a year-round average effect. In the model, the interference term acts multiplicatively on HMPV transmission and depends on normalized weekly RSV incidence, such that suppression is strongest only when RSV activity is at its seasonal peak. Because RSV circulation is seasonal and remains low during parts of the season, the time-averaged effect of RSV interference on HMPV transmissibility is modest overall. Consequently, RSV vaccination removes only a small average suppression of HMPV transmission, which explains why even substantial reductions in RSV circulation produce only modest increases in HMPV incidence. In addition, reductions in RSV-associated lower respiratory tract disease exceed reductions in RSV infection and transmission because severe RSV outcomes are concentrated in vaccine-targeted age groups.

There are limitations to our study. First, we did not have data on the genetic strains of HMPV causing LRT cases over time and across different regions. Therefore, we did not explicitly model interactions between HMPV types A and B, whose alternating activity may contribute to the biennial epidemics observed in some regions. However, these interactions are unlikely to be the primary driver of regional differences in HMPV seasonality and periodicity. Both lineages generally exhibit similar seasonal patterns, peaking in late winter to early spring in temperate regions [5]. Future studies incorporating genetic surveillance data could enhance our understanding of how lineage-specific dynamics shape regional and seasonal patterns of HMPV. Second, age information for laboratory-confirmed HMPV cases was based on ED visit data, which comes from a different data source than the weekly case counts. Testing and reporting practices differ across age groups, regions, and healthcare settings, potentially introducing biases in the observed age distribution. For example, younger children may be more likely to be tested when presenting with respiratory symptoms, while mild cases in older children or adults may go undetected. These differences could affect estimates of age-specific incidence and transmission, and may lead to under- or overestimation of the force of infection in certain age groups. In addition, while our model accurately reproduced HMPV and RSV epidemics from 2023, predictions for post-pandemic rebounds in 2021–2022 were less accurate in some regions. Other factors, beyond Google mobility data used to modify transmission rates, may also need to be considered [60].

We have been able to characterize the spatiotemporal pattern of HMPV in the US and demonstrate that vapor pressure, minimum temperature, and precipitation as well as seasonal fluctuations in PET are not key drivers with seasonal variation in the transmission of HMPV; these factors could not help explain differences in the strength of HMPV seasonality across the different regions of the US. Our analysis highlights the correlation between the seasonal RSV and HMPV epidemics and demonstrates the role of viral interference effect from RSV on HMPV transmission. Although current use of RSV interventions reduces overall RSV-related LRT cases, we predict it will have minimal impact on HMPV burden. A better understanding of the causal mechanism underpinning the viral interference effects may help predict the timing of HMPV activity across the US and further guide the development and optimal timing of interventions for HMPV.

## Materials and Methods

### Data sources

#### Laboratory tests of HMPV and RSV

Weekly data on laboratory reports of HMPV and RSV testing from 10 Health and Human Services (HHS) regions in the US, covering June 2008 to July 2025, were obtained from the National Respiratory and Enteric Virus Surveillance System (NREVSS). Positive HMPV and RSV cases were identified using three diagnostic methods: (1) antigen detection, (2) reverse transcription polymerase chain reaction (RT-PCR), and (3) viral culture.

The raw laboratory data were rescaled based on the total number of tests to account for variations in testing practices over time [11]. We first calculated a one-year moving average of the weekly number of HMPV or RSV tests (both positive and negative tests) in each region centered on each week. We then calculated a weekly scaling factor for each region equal to the average number of HMPV or RSV tests during the entire period of reporting (i.e., epidemic seasons from 2008-2025) divided by the one-year moving average. The rescaled number of HMPV- or RSV-positive tests for each region was then calculated as the reported number of positive tests multiplied by the weekly scaling factor. Here, an epidemic season is defined as starting from July of one year (ISO week 27) to June of the following year (ISO week 26).

#### Emergency department visits

We obtained aggregate data from Epic Cosmos, a large de-identified electronic health record–based dataset that covers more than half the US population, from July 2016 to June 2025. The study population included all emergency department (ED) encounters with an acute respiratory infection (ARI) diagnosis, defined using ICD-10 codes J00–J22. Within this population, we calculated the proportion of encounters with a test performed for HMPV, identified using LOINC (Logical Observation Identifiers Names and Codes) codes 60425-6, 60266-4, 77024-8, 101290-5, 40979-7, 82165-2, 92134-6, 38917-1, 67820-1, 80587-9, 67821-9, 40978-9, 89651-4, 91809-4, 92978-6, 53249-9, and 92881-2. Test positivity was defined as an abnormal result for any of these codes. Data were aggregated by state and age group (<1 year, 1–5 years, 5–18 years, 18–50 years, 50–65 years, and ≥65 years).

#### Demographic data

Information about population size in each age group was obtained from the US Census Bureau’s American Community Survey. Birth rates varied between regions and over time based on the crude annual birth rate for each HSS region from 1990 to 2019. These were obtained from https://wonder.cdc.gov/controller/datarequest/D66. To capture aging among infants and children more accurately in our mathematical model, we divided the <1 year and 1-4 years age class into 12-month age groups. The remaining population was divided into 5 age classes: 5-9 years, 10-19 years, 20-39 years, 39-60 years and >60 years old. Individuals were assumed to age exponentially into the next age class, with the rate of aging equal to the multiplicative inverse of the width of the age class.

We used the *smooth*.*spline* function (with 10 degrees of freedom) implemented in R (version 4.3.2) to interpolate weekly birth rates. We estimated the net rate of immigration/emigration for each age group (detailed in **Text S1**) to produce a rate of population growth and age structure similar to that of each region. Data on age-specific contact rates were obtained from [26] where age-specific contact patterns were estimated in the US.

#### Climatic data

Monthly climatic data were obtained from the TerraClimate dataset, which provides global, high-resolution (1/24°, ~4 km) climate data derived from the interpolation of ground-based meteorological station observations and other climate products (available from http://www.climatologylab.org/terraclimate.html). The climatic variables used were minimum temperature (°C), vapor pressure, precipitation, and potential evapotranspiration (PET). These monthly climatic variables were extracted for each US Health and Human Services (HHS) region for the period from July 2008 to June 2019 by averaging values across the entire region and were normalized to range between –1 and 1 for incorporation into the HMPV transmission model.

#### Google mobility data

Weekly mobility data were obtained from the Google Community Mobility Reports (available from https://www.google.com/covid19/mobility). These reports provide information on six categories of mobility: retail and recreation, grocery and pharmacy, parks, transit stations, workplaces, and residential. For each location, the reports provide weekly values representing the percent change in visits to locations in each category relative to pre-pandemic baseline levels, defined as the median value for the corresponding day of the week during January 3–February 6, 2020. These weekly mobility measures were aggregated for each US region for analysis.

The center of gravity and the intensity of HMPV or RSV activity The center of gravity of HMPV or RSV activity for each season and region (*G*_*s,r*_) was measured as the mean epidemic week, with each week weighted by the weekly number of positive 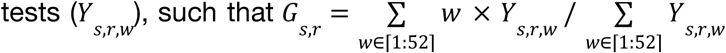, where *w* is an index for the week of each epidemic season, *s*, and region, *r*. The center of gravity is a measure of the mean epidemic week, indicating the average timing of RSV activity. This measure has been used repeatedly in the literature for RSV and other pathogens [11,61–64]. The intensity of HMPV or RSV activity for each season and region was calculated as the peak number of cases of each epidemic.

### Dynamic model description for HMPV

Here, we used an age-stratified SIS (Susceptible-Infectious-Susceptible) model for HMPV transmission dynamics. The model was proposed by Pitzer et al. [11] to study the environmental drivers of the spatiotemporal dynamics of RSV in the US. The model assumes individuals are born with protective maternal immunity, which wanes exponentially, leaving the infants susceptible to infection. We assumed a progressive build-up of immunity following up to four previous infections. Following infection with HMPV, individuals develop partial immunity, reducing the rate of subsequent infection and relative infectiousness of the following infections. We also assumed subsequent infections have a shorter recovery time compared to the primary infection. The model was described by a system of ordinary differential equations; see **Text S2** for details.

### Parameter estimation

For each region, we calibrated the model to both the weekly positive tests for HMPV from July 2008 to June 2019 and the age distribution of ED visits with HMPV positivity from 2020 to 2025. We estimated five HMPV transmission parameters: a seasonal amplitude (α), a seasonal offset (ϕ), a baseline transmission rate (β), and reporting fractions (*f*_1_ for age <18 and *f*_2_ age ≥18) to scale HMPV-related LRT infections. We estimated these parameters using a maximum likelihood approach, assuming the number of HMPV-related LRT infections during each week was Poisson distributed with a mean equal to the model-predicted number times the estimated reporting fractions; age-specific counts were jointly fit using a multinomial distribution with probabilities proportional to the model-predicted age distribution of LRT infections corrected for the differences in testing probabilities by age groups in the different regions. Other parameter values are provided in **Table S2**. We seeded the model with one HMPV-infected individual in each age group except the <1 year group then used a burn-in period of 47 years to ensure the model reached a quasi-equilibrium steady state.

For the model testing the climate hypothesis, we estimated additional parameters, including the seasonal amplitude parameters for vapor pressure (α_𝒱*p*_), minimum temperatures (α _*temp*_), precipitation (α _*pp*_), and PET (α_*pet*_). The force of infection is given by λ = β_0_ (1 + α cos(2π_𝒱_*t* − ϕ) + *a* _*climate*_*climate*)*I* ^*^, where *Climate* are normalized data of climatology variables, and denotes all infection states. Note that we tested each variable separately. For the models with viral interference from RSV, we estimated an additional parameter, η. The force of infection is given by: λ = (1 + η *RSV*)β_0_ (1 + α cos(2π𝒱*t* − ϕ))*I*^*^, where *RSV* are normalized data of RSV-related LRT cases. Rather than explicitly modeling co-infection with RSV and HMPV, we assume that RSV infections are directly proportional to the data on diagnosed RSV-related LRT cases and affect HMPV transmission to all age groups equally.

### Profile likelihood

To assess uncertainty in the viral interference parameter, we constructed a profile likelihood confidence interval. Starting from the maximum likelihood estimate (MLE), we defined a grid of fixed η values spanning a range both below and above the MLE, with a denser spacing in the neighborhood of the MLE to improve resolution. At each grid point, the remaining model parameters were re-optimized by maximizing the likelihood with η held fixed, using a multi-start strategy to reduce the risk of convergence to local optima. The 95% profile likelihood confidence interval was then defined as the set of η values for which the profile negative log-likelihood did not exceed the MLE value by more than χ^2^_1_(0.95)/2 ≈ 1.92, consistent with Wilks’ theorem.

### RSV model

To model RSV transmission dynamics, we used the same age-stratified SIS model. We calibrated the RSV model separately to only weekly RSV cases from July 2008 to June 2019. We estimated four transmission parameters: a seasonal amplitude parameter, a seasonal offset parameter, a baseline transmission rate parameter, a reporting fraction parameter to scale infections with RSV to positive cases. We estimated these parameters using a maximum likelihood approach, assuming the number of RSV cases during each week was Poisson distributed with a mean equal to the model-predicted number times the estimated reporting fraction. The model was described by a system of ordinary differential equations; see **Text S2** for details.

### Model Validation

To validate the models, we simulated HMPV and RSV epidemics following the COVID-19 pandemic. Following a previous study [18], we scaled the force of infection for both viruses by a time-varying constant, *N*(*t*), estimated from Google mobility data to capture the impact of non-pharmaceutical interventions (NPIs). The constant was computed weekly as the average percent change from baseline across retail and recreation, grocery and pharmacy, transit stations, and workplace categories. After October 15, 2022, when mobility data were no longer reported, we assumed transmission returned to baseline. Because the relationship between mobility changes and transmission reduction is uncertain, we modeled *m*(*t*) = *s*(1 + *N*(*t*)) and conducted a sensitivity analysis to test a range of scalars for each region, *s*, selecting the one that maximized the post-pandemic log-likelihood for both RSV and HMPV. Therefore, the force of infection of HMPV or RSV is expressed as λ = *s*(1 + *N*(*t*))β_0_ (1 + α cos(2π𝒱*t* − ϕ))*I* *. Notice that a negative sign for *N*(*t*) suggests a decrease in mobility. We incorporated the intervention effects for RSV during the validation period (2023–2025) and assumed an average coverage rate of 50%, based on [50,56].

### Model RSV immunization strategies

In this work, we considered three RSV intervention strategies (**Fig. S17**). To model monoclonal antibodies for infants, we assumed that immunized infants have prolonged immunity against RSV infection. This framework was proposed in a previous modeling study [65]. Efficacy is determined by the waning rate of prophylaxis, with an average duration of protection of 275 days [65–67]. After this protection wanes, immunized infants become susceptible to RSV infection and have the same risk of infection as unimmunized infants. To model maternal vaccination, we assumed that successful maternal vaccination increases the level of transplacentally acquired immunity in infants born to vaccinated mothers, thereby reducing both the risk of RSV infection and developing lower respiratory tract infection (LRTI). The mean relative risk reduction was based on vaccine efficacy estimates from the Phase 3 MATISSE (Maternal Immunization Study for Safety and Efficacy) trial of bivalent RSV pre-fusion F protein vaccine (RSVpreF), which reported 82.4% (95% CI: 57.5–93.9%) efficacy against severe medically attended RSV-associated LRTI within 90 days of birth, and 70.0% (95% CI: 50.6–82.5%) within 180 days of birth [68]. These findings were consistent with the prespecified interim analysis of the same trial [52], which demonstrated 81.8% (99.5% CI: 40.6–96.3%) efficacy against severe RSV-associated LRTI within 90 days. The duration of vaccine-induced protection was assumed to be 5 months (150 days), consistent with the observed decline in vaccine efficacy between 90 and 180 days in the MATISSE trial, and substantially longer than the approximately one month over which naturally acquired transplacental immunity is estimated to persist. To model older-adult vaccination, we assumed that vaccination reduces the risk of infection and the risk of developing LRTI. All parameters values are provided in **Table S5**, and see **Text S3** for detailed model equations. Notably, none of the three interventions were assumed to reduce infectiousness upon breakthrough infection; immunized individuals who subsequently acquired RSV were assumed to transmit at the same rate as unimmunized infected individuals. The reductions in RSV incidence observed in age groups not directly targeted by immunization therefore reflect an indirect population-level effect: by reducing the number of infectious individuals in the population, these interventions lower the overall force of infection experienced across all age groups. We varied the coverage rate of all interventions from 0% to 100% over 2023–2027. The change in RSV- or HMPV-related LRT infection burden was calculated by aggregating all cases from 2025 to 2027 and comparing them to the baseline scenario (i.e., coverage rate = 0).

## Data Availability

All data produced in the present work are contained in the manuscript

## Data and code availability

We used the R statistical software (v4.3.2) for all statistical analyses and visualization. The laboratory data is available by contacting the NREVSS coordinator at the Centers for Disease Control and Prevention. The HMPV data from Epic Cosmos and code used in this study is publicly available on Github: https://github.com/keli5734/hmpv_seasonality. Figures 2A, 2C, 3C, 3D and 3E were generated in R using U.S. Census TIGER/Line shapefiles accessed through the tigris R package. The base map shapefiles are publicly available from the U.S. Census Bureau at TIGER/Line Shapefiles. Terms of use and licensing information are available at U.S. Census Bureau Terms of Service. These data are publicly available and compatible with the CC BY 4.0 license.

## Acknowledgments

This work was partially supported by a grant from the National Institutes of Health (R01AI137093) awarded to D.M.W and V.E.P. This work was additionally partially supported by the Merck Investigator Studies Program contracts 103595 (N.D.G, V.E.P, D.M.W) and 103622 (V.E.P, D.M.W, N.D.G). The content is solely the responsibility of the authors and does not necessarily represent the official views of the National Institutes of Health. Data used in this study came from research performed using Epic Cosmos, a dataset created in collaboration with a community of health systems using Epic representing more than 300 million patient records from over 1700 hospitals and 40000 clinics as of September 2025. The community represents patients from all 50 states, D.C., Canada, Lebanon, and Saudi Arabia. We also thank all the participating NREVSS laboratories for their contribution to the data analyzed in this manuscript.

## Author Contributions Statement

K.L. conducted the analyses and wrote both the initial and revised manuscripts. S.P. and J.K. extracted the data for analysis. N.D.G provided content expertise, supervision and funding acquisition. D.M.W. and V.E.P. provided content expertise, supervision, and oversaw the analyses for both the initial and revised manuscripts. All authors contributed to drafting and approved the final version of the manuscript.

